# Human-Centered Design of an Artificial Intelligence (AI) Monitoring System: The Vanderbilt Algorithmovigilance Monitoring and Operations System (VAMOS)

**DOI:** 10.1101/2025.06.05.25329034

**Authors:** Megan E. Salwei, Sharon E. Davis, Carrie Reale, Laurie L. Novak, Colin G. Walsh, Russ Beebe, Scott Nelson, Sameer Sundrani, Susannah Rose, Adam Wright, Michael Ripperger, Peter Shave, Peter Embí

## Abstract

**Background.:** As the use of AI in healthcare is rapidly expanding, there is also growing recognition of the need for ongoing monitoring of AI after implementation, called *algorithmovigilance*. Yet, there remain few systems that support systematic monitoring and governance of AI used across a health system. In this study, we describe the human-centered design (HCD) process used to develop the Vanderbilt Algorithmovigilance Monitoring and Operations System (VAMOS).

**Methods.:** We assembled a multidisciplinary team to plan AI monitoring and governance at VUMC. We then conducted nine participatory design sessions with diverse stakeholders to develop prototypes of VAMOS. Once we had a working prototype, we conducted eight formative design interviews with key stakeholders to gather feedback on the system. We analyzed the interviews using a rapid qualitative analysis approach and revised the mock-ups. We then conducted a multidisciplinary heuristic evaluation to identify further improvements to the tool.

**Results.:** Through an iterative, HCD process, we identified key components needed in AI monitoring systems. We identified specific data views and functionality required by end users across several user interfaces including a performance monitoring dashboard, accordion snapshots, and model-specific pages. We distilled general design guidelines for systems to support AI monitoring throughout its lifecycle. One important consideration is how to support teams of health system leaders, clinical experts, and technical personnel that are distributed across the organization as they monitor and respond to algorithm deterioration.

**Conclusion.:** VAMOS enables systematic and proactive monitoring of AI tools in healthcare organizations. Our findings and recommendations can support the design of AI monitoring systems to support health systems, improve quality of care and ensure patient safety.

## INTRODUCTION

Artificial intelligence (AI) is becoming increasingly prevalent in healthcare, with the potential to transform healthcare delivery by improving clinician well-being and patient safety while reducing healthcare costs [1]. AI is being implemented across the continuum of care to assist with medical imaging [2], notetaking during clinical encounters [3], responding to in-basket messages [4], and diagnosing high-risk conditions such as sepsis [5], suicide [6], and post-partum hemorrhage [7]. Yet, despite the potential benefits, AI introduces several well-documented risks [8, 9]. One major concern is systemic issues with the data embedded in AI that can further exacerbate differences in how well models work for groups of people [10]. Another concern is the deterioration of AI accuracy over time due to changes in patient populations, policies, and/or how we capture data [11–13]. Performance can also evolve as implemented models change behavior [14]. Moreover, end-user interactions with AI and the broader sociotechnical system - including clinical workflow and the physical and organizational environment - can also significantly affect AI performance, use, and impact [15–17]. Additionally, for legal and ethical reasons, effective AI monitoring is needed to avoid the risk of harm to patients and other individuals [18–21]. These issues provide the impetus to routinely monitor AI models and their use after implementation to ensure patient safety and impact sustainability.

Inspired by the concept of pharmacovigilance, Embí [22] proposed the need for *algorithmovigilance*, defined as “the scientific methods and activities relating to the evaluation, monitoring, understanding, and prevention of adverse effects of algorithms in health care.” (pg. 2). Across national organizations and the informatics community there have been growing calls for ongoing and continuous monitoring of algorithms after implementation [23–26]. Some organizations have leveraged clinical AI governing boards [27], algorithmic auditing [28, 29], and tools for localizing sources of errors in models [30] as a means of algorithmovigilance. Several frameworks have been proposed to guide these activities [27, 29, 31]. Davis et al. [12] described a continuum of algorithmovigilance approaches: reactive (end-user reporting), preventative (stability-focused design), preemptive (technical oversight), and responsive (data-driven oversight). Yet, AI monitoring to date largely relies on reactive rather than proactive approaches, and effective identification and monitoring of AI tools remains a challenge even in well-resourced health systems [32].

Several tools have been developed to support more proactive and robust AI monitoring [33, 34]. For instance, Epic’s Seismometer is one promising open-source tool that supports performance and fairness monitoring of predictive models [35]. Yet, most existing monitoring systems focus on the needs of data scientists rather than organizational leaders, clinical stakeholders and patients, and few systems support collaboration between clinical and technical teams to investigate and restore algorithmic deteriorations. Additionally, there remains limited research on end-user perspectives and needs for these systems. New systems are needed to support comprehensive and systematic monitoring AI in healthcare [13, 22]. In this study, we developed a new technology to support systematic AI monitoring in one health system.

Human-centered design (HCD) is a human factors engineering approach that engages end-users throughout iterative cycles of design to develop highly usable interactive systems [36, 37]. The gold standard for systems design across industries [36, 38, 39], HCD follows four primary steps: (1) understand and specify the context of use, (2) specify user requirements, (3) produce design solutions to meet user requirements, and (4) evaluate the designs against the requirements [36]. Typically, an HCD process starts with studying the environment of use and intended users to understand their workflow and needs. However, this process is limited when designing a system for a work domain that does not yet exist, referred to as the “envisioned world problem” [40, 41]. This is the case for AI monitoring systems. Entirely new departments and roles will likely be developed in the coming years to support the ongoing maintenance and monitoring of AI models within healthcare systems. Miller and Feigh [42] describe two specific pathways for designing for the envisioned world: (1) the technology-driven path and (2) the work-driven path. The technology-driven path focuses first on the technology’s capabilities, building the technology, and then introducing it into the work domain. This approach largely ignores the broader sociotechnical system and its impact on the technology’s use. In contrast, the work-driven path starts by focusing on the context of future work and the tasks needed to accomplish work goals. This (superior) approach is more likely to result in technologies and systems that support end-user needs. To do this, early participatory design and iterative evaluation with relevant stakeholders is needed [41].

In this study, we describe our work-driven, HCD approach used to develop a novel algorithmovigilance system - the Vanderbilt Algorithmovigilance Monitoring and Operations System (VAMOS). We propose generalizable design recommendations for developing algorithmovigilance programs to support AI monitoring across institutions.

## METHODS

We leveraged an HCD approach to develop VAMOS (Figure 1 and Table 1). This study is part of a larger project focused on the safe implementation and use of AI in healthcare [43]. This study was approved by the Vanderbilt University Medical Center (VUMC) Institutional Review Board.

**Figure 1.**
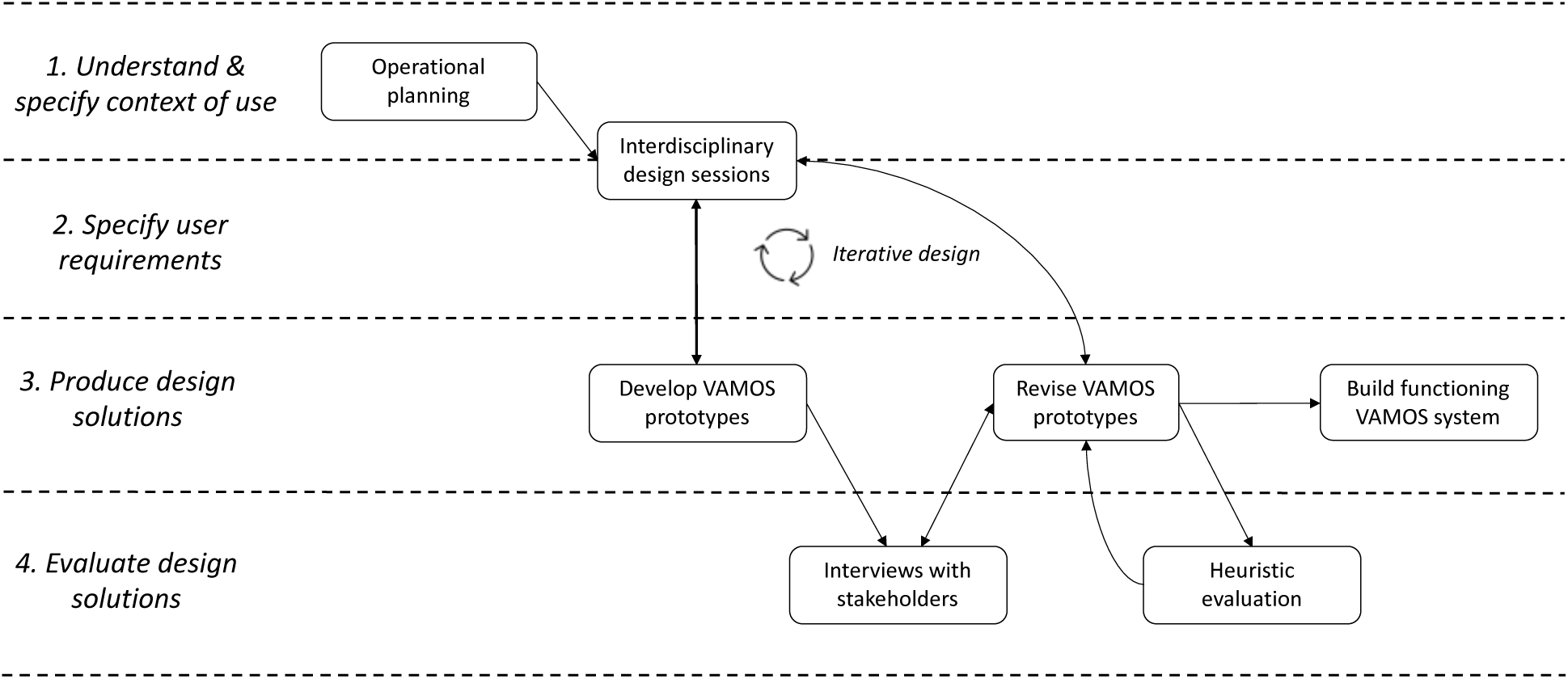
Overview of human-centered design process to develop VAMOS

**Table 1.**
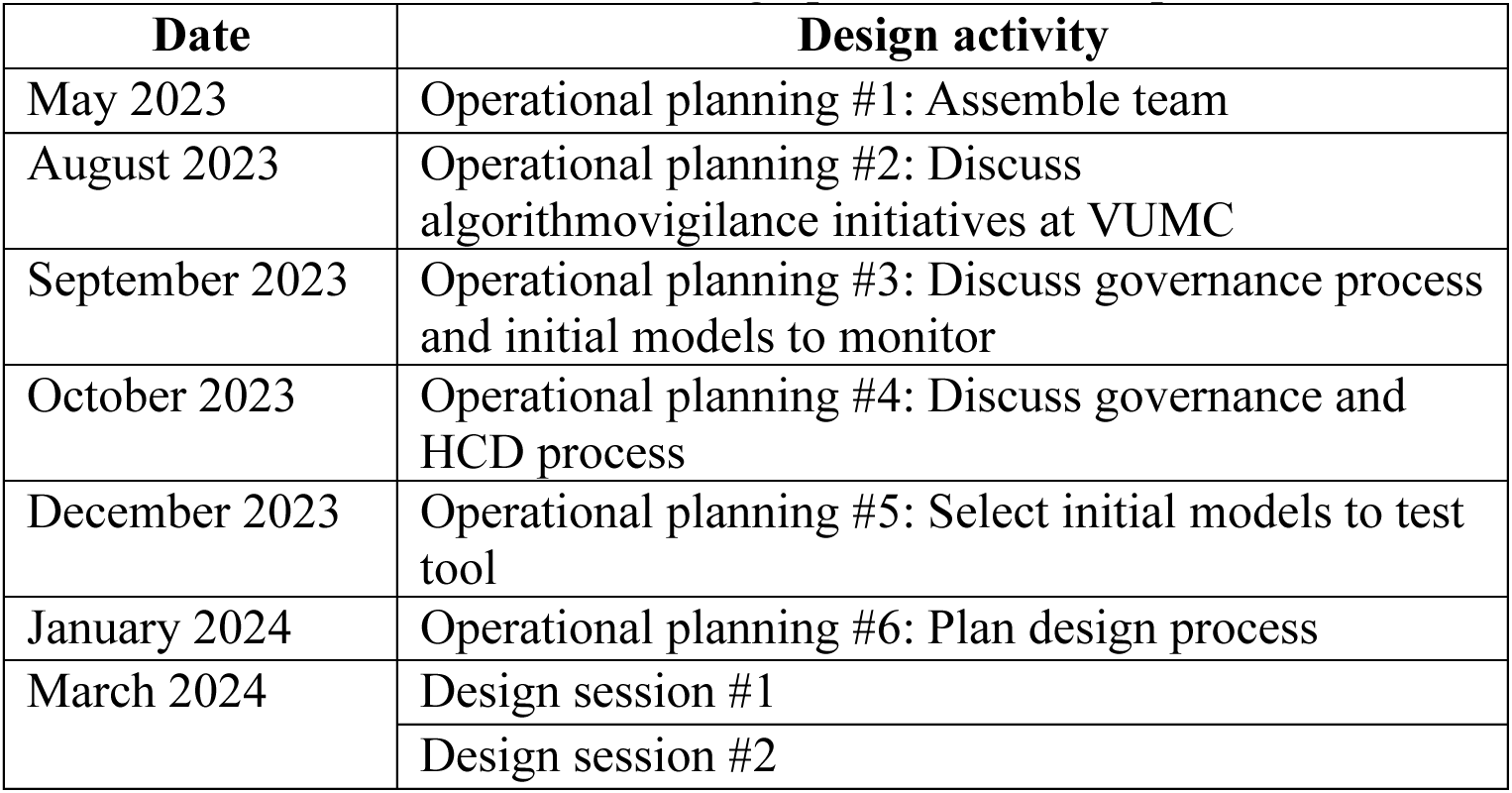
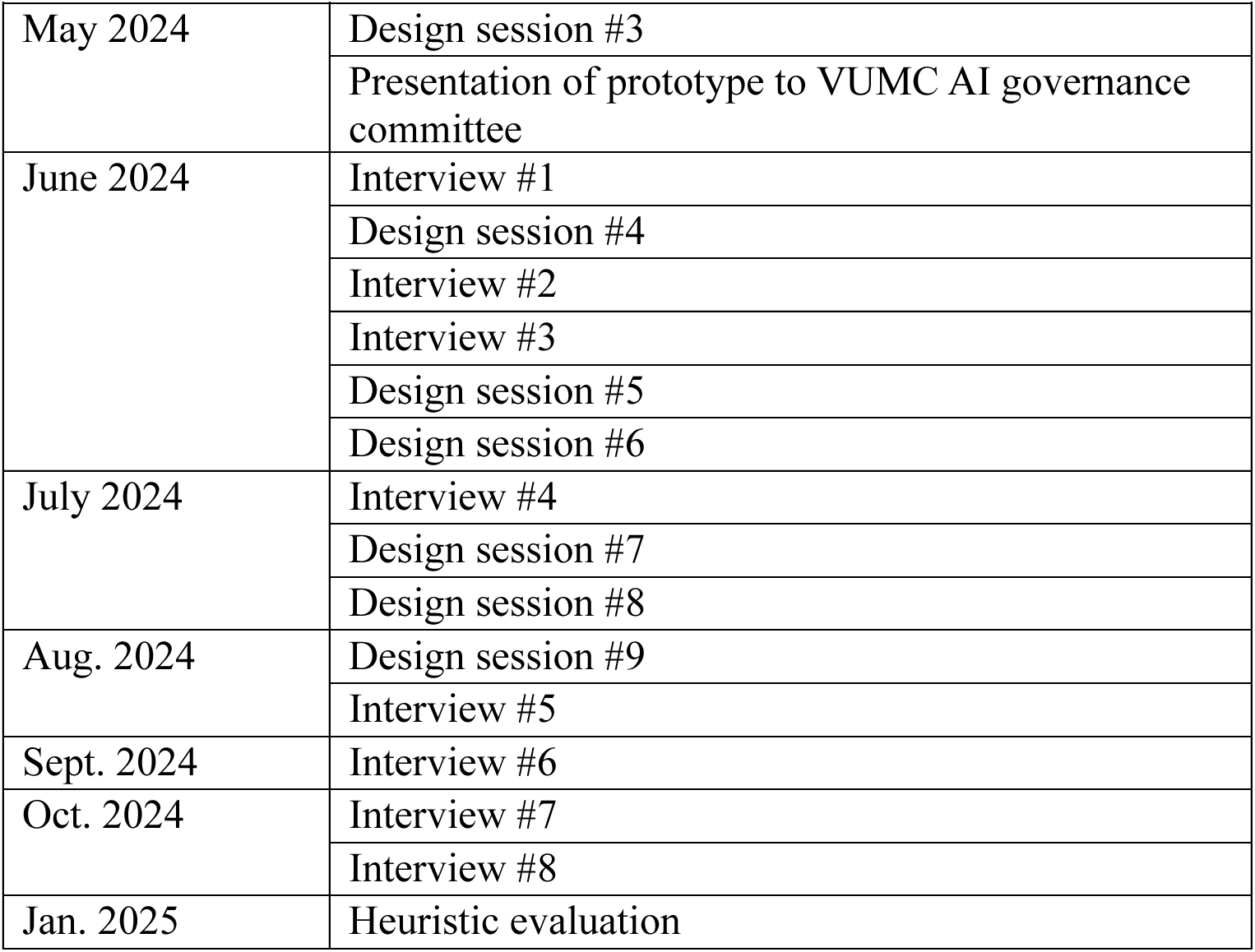
Human-centered design process to develop VAMOS

### Operational planning for AI governance and monitoring

In 2023, we assembled a multidisciplinary team to discuss AI monitoring and governance at VUMC. We engaged experts in AI modeling and evaluation, health equity, privacy and security, clinical decision support implementation, health IT, organizational design, and human factors engineering. This collaboration focused on initial goals and concepts for algorithmovigilance and the necessary infrastructure to support AI monitoring. We discussed current VUMC AI activities including the use of both generative and analytic AI, as well as existing functionality and algorithms currently deployed in the EHR for monitoring. Over the next 7 months, we continued to meet regularly and planned the development of a prototype system for AI monitoring at VUMC. Each planning session included between 4-11 people and lasted an average of 51 minutes. Throughout these sessions, we drafted a governance process, identified candidate models to include in the early prototype, and planned the HCD process.

### Participatory design sessions

Starting in 2024, we designed the user interfaces for the VAMOS system. Each design session included diverse stakeholders including experts in biomedical informatics, health IT, human factors engineering, ethics and governance, and graphic design. These sessions started by focusing on three questions, in line with the work-driven path described by Miller and Feigh [42]: (1) *who are the end users* of the tool, (2) *what are their goals and information needs*, and (3) *what tasks need to be accomplished with the system*. We used Microsoft PowerPoint to develop mock-ups of the tool, which we iteratively refined throughout the design sessions. We engaged with hospital leaders involved in the AI governance process to ensure our tool aligned with the current and envisioned processes (e.g., the intake process for newly developed algorithms). We conducted 9 design sessions, each lasting between 1-2 hours (total 10.5 hours) and including 5-11 people.

### Design interviews

Once we developed an initial prototype of VAMOS, we conducted formative design interviews with key stakeholders. We interviewed experts in AI development and implementation, AI ethics and governance, CDS monitoring, AI privacy and security, hospital informatics leadership, and those who respond to breakdowns in current IT systems across the health system. Because the actual end-users of VAMOS are not yet defined, we identified people in existing roles that use similar skills as those that will be needed for VAMOS (e.g., CDS monitoring, identifying anomalies, responding to health IT breakdowns), an approach used in prior studies designing for the envisioned world [44]. One or two human factors researchers conducted each interview over Microsoft Teams. We started each interview by asking the interviewee to describe their role, responsibilities, and involvement with AI development and implementation. We then asked about any current monitoring activities they perform in their role relating to AI or other health IT tools.

If applicable, participants shared their screen and displayed monitoring systems that they currently use. We then asked participants about the information they would want for monitoring AI models across the organization. In the second half of the interview, we elicited specific feedback on the VAMOS prototypes. The interviewer displayed the prototypes and gathered participant feedback on each prototype, asking probing questions to understand if any information was unclear. All interviews were audio-recorded and transcribed. After conducting four interviews, we revised the mock-ups to incorporate participant feedback. We then continued interviews with additional stakeholders to identify further refinements to the tool. Two human factors researchers coded the interviews to identify design modifications using a rapid qualitative analysis approach [45]. We created a grid with one row for each interview with columns to collect the interviewee’s role, prior experience with AI development, implementation, or monitoring, their information needs for AI monitoring systems in general, and their specific feedback on the VAMOS prototypes. Two human factors researchers reviewed the interview transcripts and systematically completed the grid, identifying common themes requiring design modifications to VAMOS. We conducted 8 interviews, which lasted an average of 49 minutes (total: 6.6 hours).

### Heuristic evaluation

We then conducted a 1-hour group-based heuristic evaluation with a multidisciplinary team including experts in human factors, organizational design, and biomedical informatics (e.g., AI performance monitoring, AI predictive modeling). All participants in the heuristic evaluation were sent the current VAMOS prototypes and a list of human factors design principles to review prior to the meeting. We used the design principles (i.e., Ergonomic Criteria) of Scapin & Bastien, [46], with addition of some specific examples from the Nielsen-Schneiderman heuristics [47]. To ensure we were considering the integration of VAMOS in user workflows, we also used the Checklist for Workflow Integration developed by Salwei et al., [48], which we slightly tailored for the user case and heuristic evaluation approach.

In the heuristic evaluation session, we started by displaying the 3 prototypes and allowing everyone about five minutes to write down any design principle violations or points of confusion in the interfaces. We then spent the rest of the session sharing the issues identified and discussing approaches to resolve them. The heuristic evaluation was audio-recorded and transcribed for analysis. One human factors researcher reviewed the transcript to identify each design principle violation discussed by the group along with the recommendations to resolve it. We revised the prototypes based on the findings.

## RESULTS

Through the iterative design process, we developed and refined VAMOS. Here, we report on our findings concerning the three primary user interfaces: (1) the performance dashboard (Figure 2), (2) the accordion snapshot (Figure 3), and (3) the full model detail page (Figure 4). Our findings shed light on the general functionality needed for algorithmovigilance systems across healthcare organizations.

**Figure 2.**
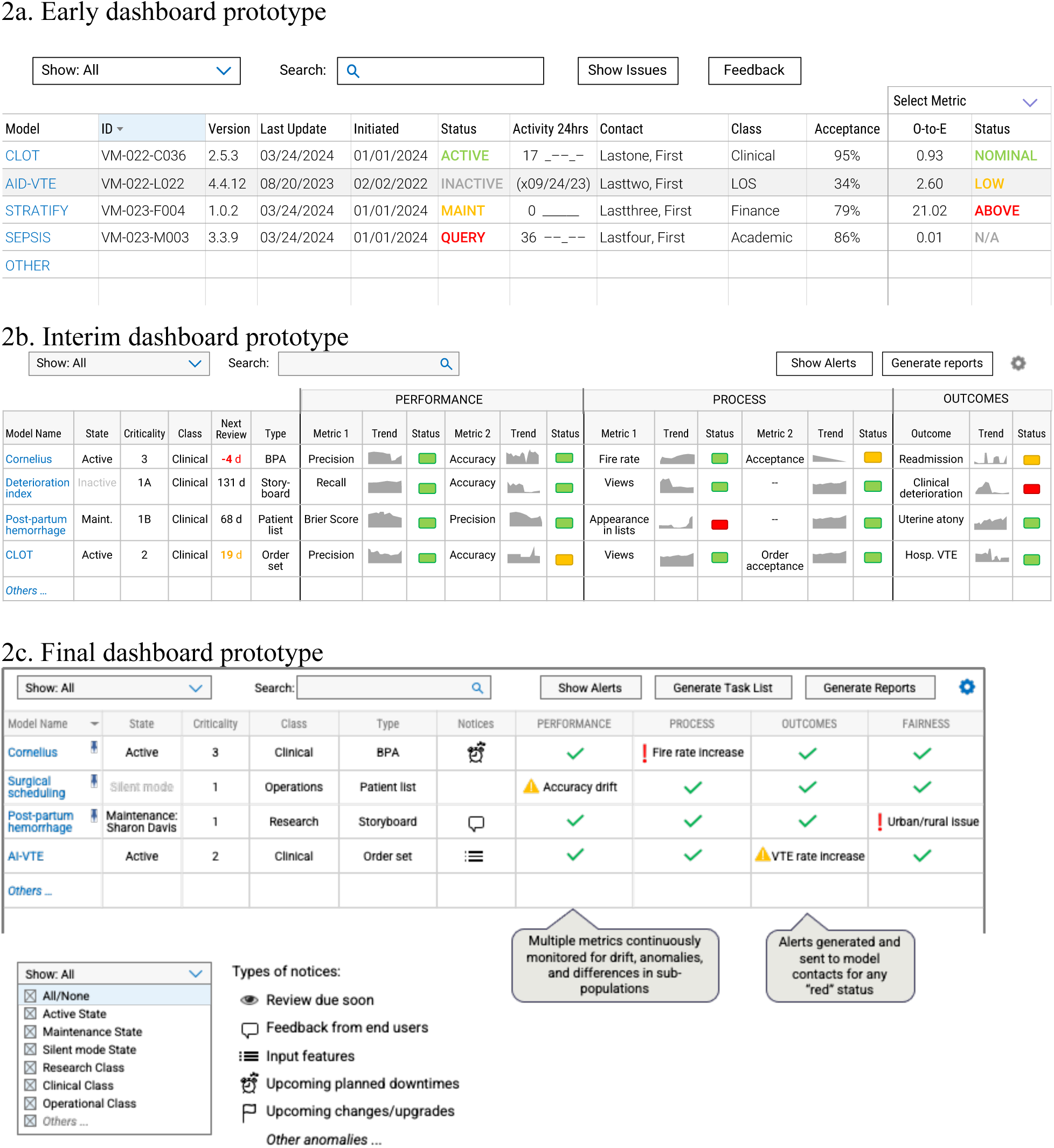
Algorithmovigilance performance dashboard (early mock-up on top, interim mock-up in the middle, final mock-up on the bottom)

**Figure 3.**
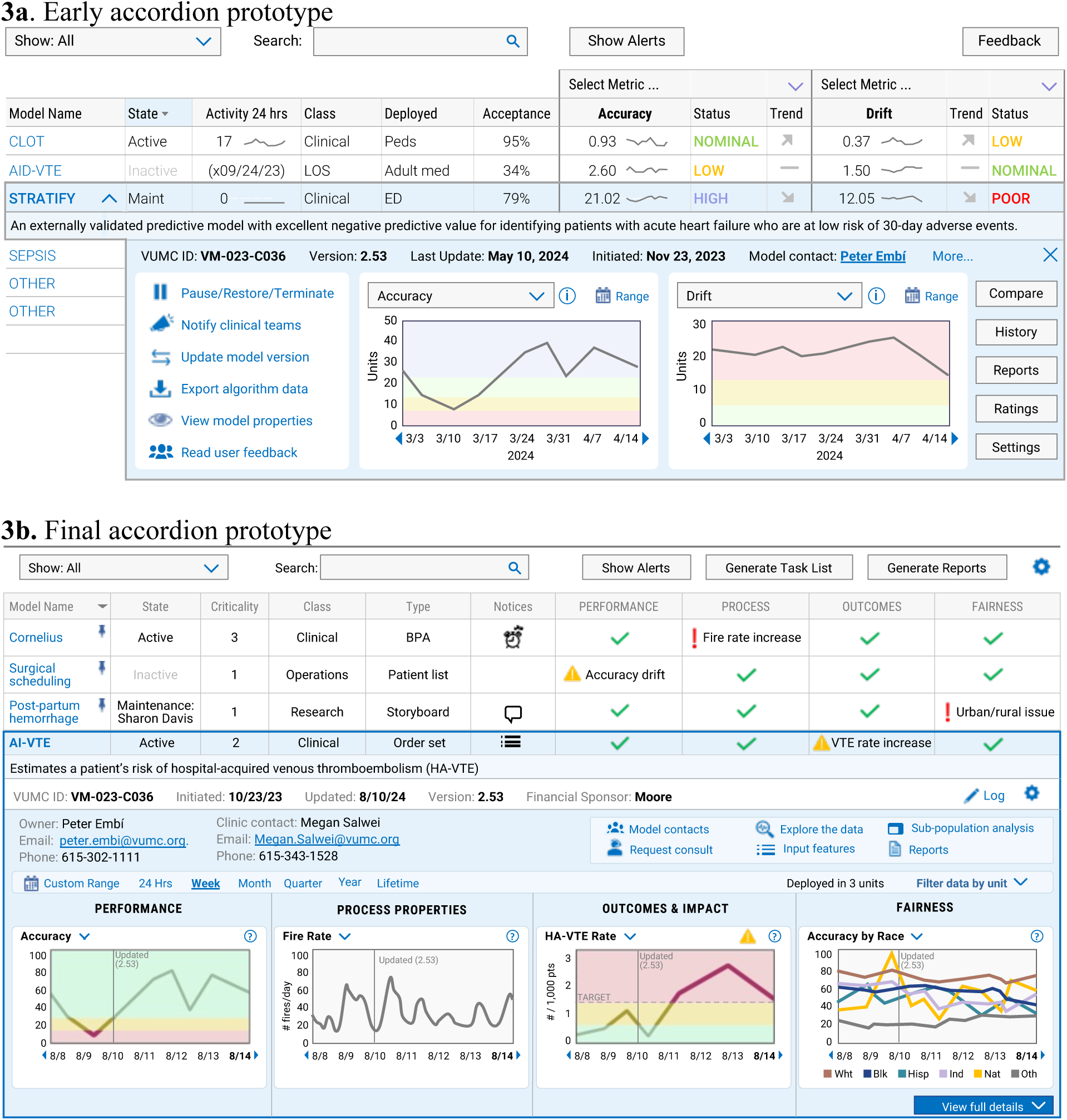
Algorithmovigilance accordion view (early mock-up on top, final mock-up on bottom)

**Figure 4.**
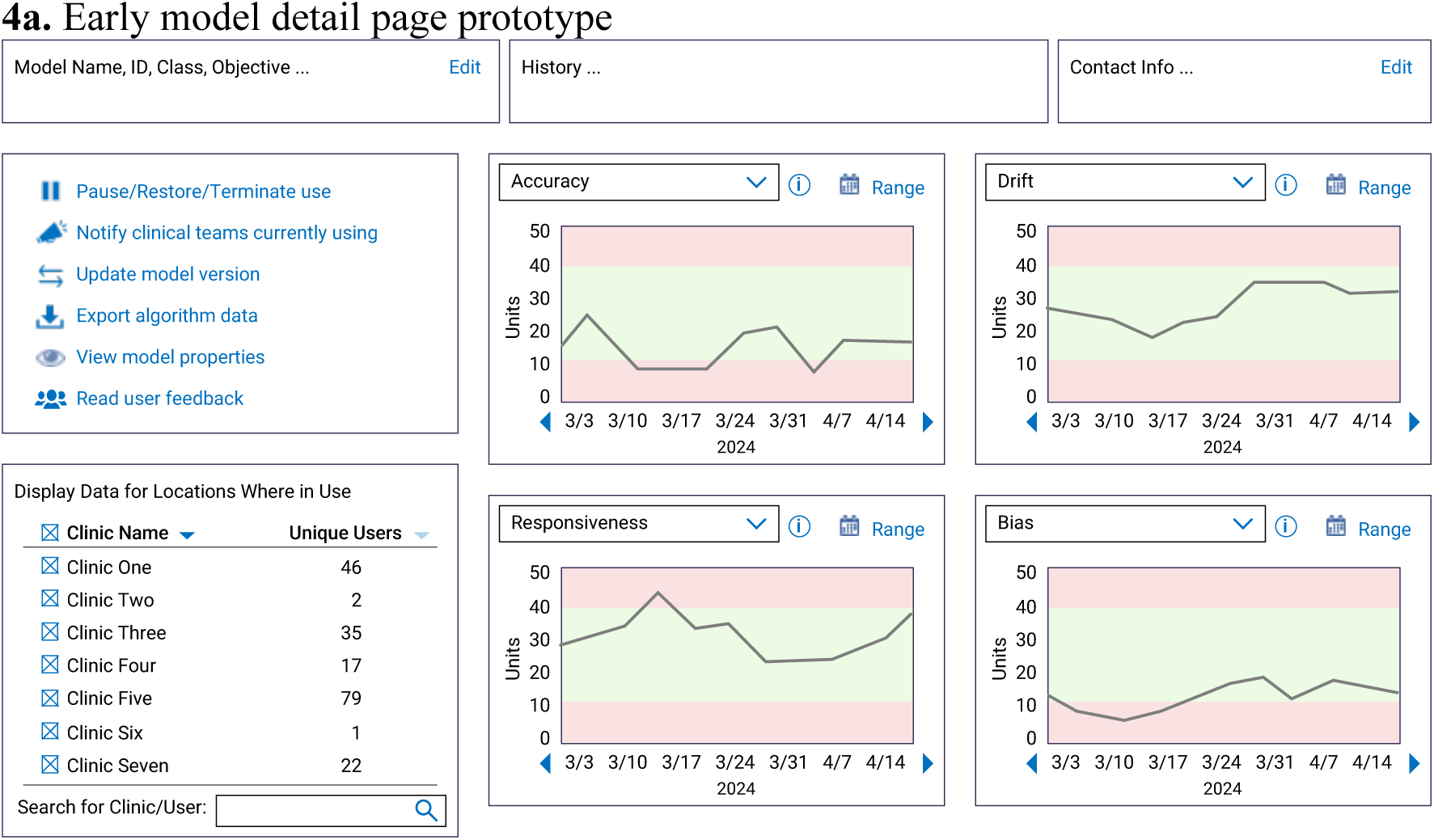
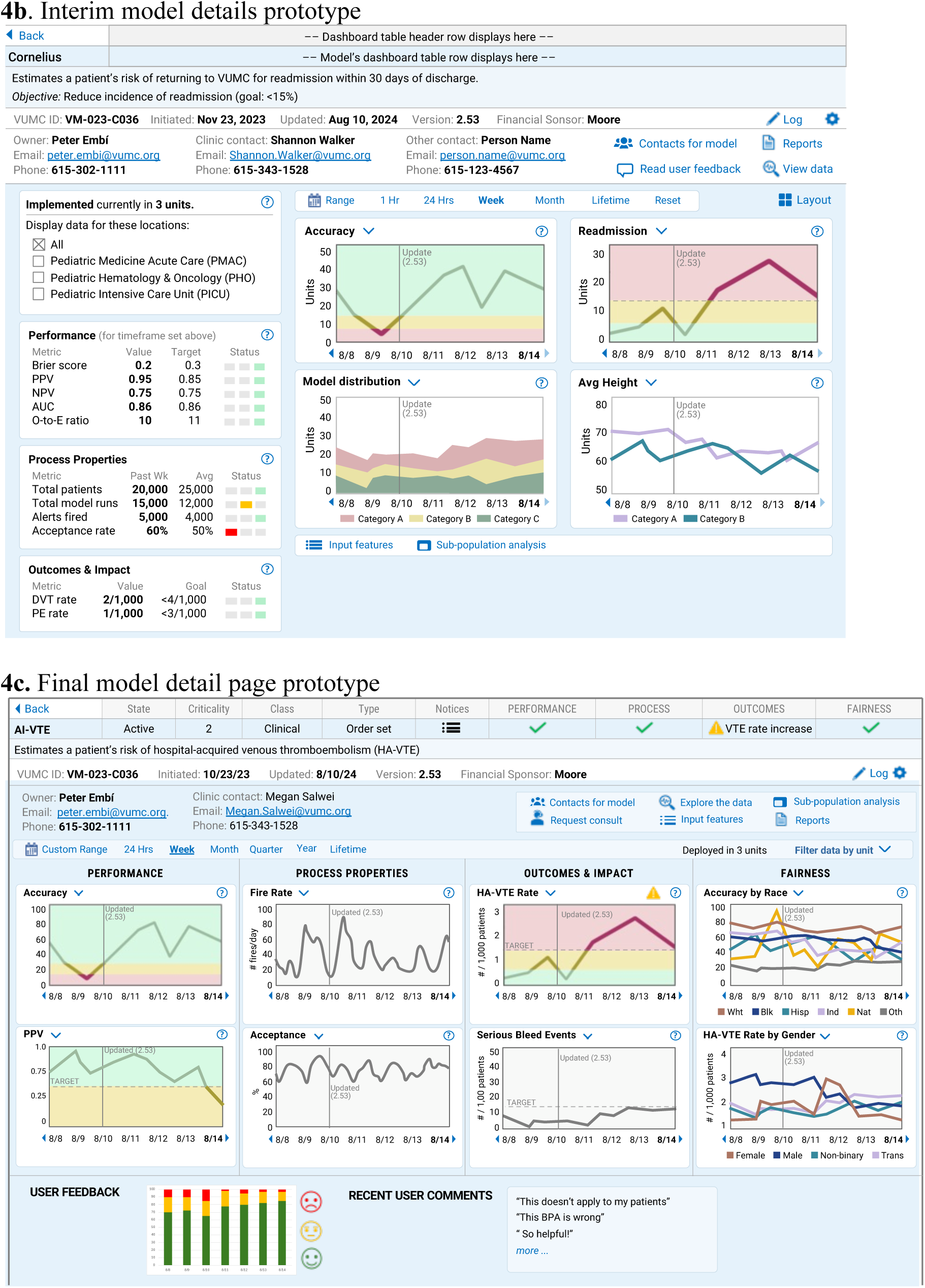
Algorithmovigilance model detail page (early mock-up on top, interim mock-up in the middle, final mock-up on the bottom)

### Performance dashboard

The *performance dashboard* is the go-to page for AI system monitoring, providing an overview of all algorithms implemented in the health system and key indicators of model health (Figure 2). The dashboard starts by listing each model’s name, state (e.g., active, under maintenance), criticality (the potential risks if the model stops working or drifts), class (i.e., clinical, operational), and type (i.e., how the model is delivered to end users). Next, any important notices for the model are shown, such as problems with input features, new end-user feedback, and any upcoming downtimes or upgrades that may affect the model. The next section of the dashboard focuses on model health across four categories: performance, process, outcomes, and fairness.

Throughout the design process, we iteratively revised the information in the dashboard based on user feedback. The main takeaway from this process was the need to minimize the information in the display, enabling users to quickly identify which models need attention. Our mock-ups initially only displayed one performance metric (see Figure 2a), which could be toggled to select different metrics (e.g., O-to-E vs PPV). During design, we found that users preferred differing metrics (e.g., accuracy, precision) depending on the use case and model type and that multiple different performance metrics should be tracked at one time. To address this, an interim version of the mock-up displayed multiple performance and process metrics for each model (see Figure 2b). Based on user feedback, we further refined the dashboard to streamline the information and minimize cognitive burden. We removed the detailed text on each model’s performance metrics and its value and instead used color (green, yellow, red) and icons (checkmark, caution sign, exclamation point) to quickly communicate the status of key metrics. One participant explained: “I don’t care if one was precision and the other one was recall… I just need you to tell me when something’s not working” (P8). All of the information in VAMOS is sourced from the VAMOS analytics suite, which stores model history and calculates metrics for each interface (e.g., accuracy, fire rate). Any anomalies (e.g., sudden increase or decrease, drift over time) with a model’s performance, process, outcome, or fairness metrics would be flagged the dashboard and additionally an alert would be sent directly to the model owner.

We also received feedback on the types of information to include in VAMOS. Early mock-ups primarily focused on performance information (e.g., model accuracy). During the design interviews, participants emphasized the importance of seeing process information (e.g., model run rate, user acceptance) and the clinical impact of models (e.g., outcomes), with one application developer explaining, “Ultimately, what we really care about is did it solve the problem it was intended to solve?” (Participant 3). We expanded the dashboard to include four main categories: performance, process, outcomes, and fairness.

The adaptability of the interface was also an important design consideration. Due to the broad range of users that will employ the dashboard (e.g., health system executives, data scientists, unit managers), the design team frequently discussed the need to sort and filter the dashboard. The dashboard can be sorted and filtered based on any category in the dashboard (e.g., state, class) as well as by clinical department. The “show alerts” button filters the dashboard to only show models that need attention due to new notices or issues with performance, processes, outcomes, or fairness. We also included quick definitions of key terms in the dashboard. By hovering over a header, users can access a brief description of that item (e.g., criticality). Similarly, a brief description of the model appears when hovering over the model’s name. In the heuristic evaluation, we identified several issues relating to the support of the maintenance team workflow. For instance, participants identified the need to know who was currently assigned to a model that was under maintenance. We added the name of the data scientist under the “state” column for models in maintenance.

### Accordion snapshots

The *accordion snapshot* (Figure 3) builds off of the performance dashboard by allowing users to quickly drill down into specific model information. From the performance dashboard, users can click on any model to get additional details on the model. The accordion snapshot displays a brief description of the model goals (e.g., reduce incidence of hospital-acquired venous thromboembolism), implementation date, last update date, and where the model is deployed. In line with the categories on the performance dashboard, it displays four graphs showing trends in model performance, process measures, outcomes, and fairness. These graphs indicate the timing of prior model updates (vertical line) and can be viewed over different time horizons (e.g., model lifetime, last week). During the interviews, we found that having quick access to multiple contacts for the model was an important feature. One participant described, “[if] I’m a model reviewer, project manager, and holy camoly. Like there’s something really going wrong right now…this is on, you know, this floor, and this could affect patients…you want that to be a quick decision point over who the heck they call” (P4). The top of the accordion displays the model owner and clinical contact; additional model contacts (e.g., technical personnel) are accessible from the “contacts for model” button. Users can also generate reports, explore model data, and read end-user feedback. Over the design process, we learned that users wanted a feature to support note-taking during AI governance meetings; the “log” feature on the accordion can be used to quickly take notes on necessary actions (e.g., maintenance) needed for specific models.

In the heuristic evaluation, we found that the “log” feature would also be helpful for tracking prior issues and updates made to the model. Many of the issues in the heuristic evaluation related to the design principle, “flow of tasks”, specifically, how to better support the next steps that users would have to take after viewing the information in VAMOS. We added the “request a consultation” button to help initiation of a model review by our health IT, data science, and human factors experts (i.e., the VAMOS consultative service).

### Model detail page

The *model detail* page expands on the accordion snapshot, providing a deep dive into model health. Building on the accordion snapshot, the model detail page provides additional graphs of model health metrics, for example, additional outcome measures of interest like serious bleed events for a risk score recommending VTE prophylaxis in high-risk patients. One interviewee described, “More information is better than less information. I know maybe that’s not typical in design, but like if you’re serious about monitoring these things, you want as much information as possible.” A user can click on these graphs to expand their view. They can also switch which metrics are shown on the graphs. The model detail page also gives a snapshot into user feedback, for example, if a clinician thinks an AI-based best practice advisory (BPA) is incorrect for their patient.

Overall, the VAMOS displays provide a progressive level of detail to support the tasks of model reviewers, “I don’t think that the dashboard necessarily needs to tell you what to do. But the person reviewing it has to be able to understand the information quickly enough, prioritize it fast, and be able to drill down and to find solutions”. We found that report generation is an important functionality for algorithmovigilance systems. In VAMOS, reports can be generated from both the accordion snapshot and the model detail page. Reports can be generated and tailored for specific units (e.g., on all the models in use in the hospital) or for specific model owners. Institution-wide reports can also be generated to support governance committee review and, perhaps in the future, auditing.

## DISCUSSION

In this study, we designed a novel algorithmovigilance system to support monitoring of AI algorithms across a health system. Despite the well-documented risks of AI [49], including performance degradation over time [13, 50], there are few systems to support the ongoing monitoring of AI in healthcare [33–35]. Through an iterative HCD process, we developed functional prototypes for VAMOS, which supports various algorithmovigilance activities including technical and data-driven oversight and end-user reporting [12]. We identified end-user needs for AI monitoring in healthcare and outlined how to support these needs through a performance dashboard, accordion snapshots, and model-specific pages. To our knowledge, this is the first study that has solicited end-user feedback on algorithmovigilance systems. While this study was conducted in one academic medical center, our findings can inform the design of algorithmovigilance systems more broadly in healthcare. Development and use of an AI monitoring system will be essential for any health system aiming to use AI safely, and it is likely that tracking and reporting on AI performance, process, and outcome metrics, such as those described in VAMOS, will be required by regulatory agencies (e.g., The Joint Commission) in the future. The work reported in this paper provides one example of how these data may be presented and how organizations can approach AI monitoring activities in practice. Rather than prescribing *how* organizations should go about AI monitoring, we provide insights into the information and activities that need to be supported for ongoing AI monitoring.

We identified several design requirements for algorithmovigilance systems, as shown in Table 2. These findings complement existing recommendations and frameworks for AI monitoring and governance more broadly [12, 25, 27, 29, 31]. These guidelines provide insights from this work that other health systems can leverage when deploying AI monitoring solutions. One important requirement is the ability to filter and sort dashboard information. A diverse range of users will likely interact with AI monitoring systems, such as health system executives, clinic managers, and data scientists. Each of these users will have varying levels of AI competencies [51], expertise, knowledge, and specific goals when engaging with the system, necessitating a flexible interface to accommodate these differences. Therefore, the ability to easily filter and sort information ensures that each user can efficiently access the most relevant data for their role, whether they are involved in research, governance, cybersecurity, or patient experience. We also included easy-to-access definitions of terms used throughout the system to accommodate different AI competencies of end users [51]. This allows the system to support the unique needs and skills of all stakeholders involved in AI monitoring and governance.

**Table 2.** Design requirements for AI Monitoring Systems

| Design requirement | Specific design features in VAMOS |
| --- | --- |
| Support a broad range of users with differing goals, expertise, and background knowledge | • Filtering and sorting of information<br>• Easily accessible definitions of terms and models<br>• Progressive level of detail in model information, e.g.,<br>○ Ability to view input features of a model, if desired<br>○ Ability to view detailed data on models, if desired |
| Support communication when model issues are identified | • Quick access to a page containing all of the contacts for a model (e.g., clinical leader, health IT management)<br>• Ability to request a consult for the accordion and model detail pages |
| Receive and display feedback from end-users | • Section of model-specific page to view end user feedback<br>• Notice on the accordion for new issues with end user feedback |
| Track notes and action items on specific models during governance committee meetings | • Log functionality in accordion dashboard |
| View prior model maintenance and updates made to a model | • Log functionality in accordion dashboard |
| Provide oversight for unit managers, governance committee meetings, and regulators | • Quickly generate reports that can be filtered for specific users, units, settings, etc. |
| Identify drift in metrics over time | • Show metrics in graph form<br>• Allow viewing of graphs across different units |
| Display trends over variable time horizons (e.g., before model implementation) | • Ability to adjust the time horizon on all graphs |
| Quickly identify models with issues | • “Notices” section of the dashboard flags any issues<br>• Red, yellow, green icons to quickly communicate model health<br>• Anomaly detection and alerts for any significant changes |
| Identify issues with input data or key features | • Can view trend information for different features (e.g., patient age) in the model details page<br>• “Notices” flag for input feature issues |
| Tailor metrics for each specific model | • Performance, process, and outcome measures are different for each model and selected at intake |
| Provide insight on all aspects of model health including performance, process, outcome, and fairness measures | • Dashboard, accordion, and model detail page have specific sections for performance, process, outcome, and fairness measures |
| Specify the goal of each model | • Definition of model in the accordion snapshot and model detail page |
| Ability to prioritize model issues that need review | • “Criticality” value to indicate the safety or organizational risk level of a model<br>• “Generate task list” button quickly sorts the dashboard according to model criticality and the current problem with the model (e.g., large vs. small model drift) |

The development of the algorithmovigilance system presented a unique design challenge. Typically, a HCD process starts with studying the environment of use and intended users for the system. However, this is not possible when designing a system for an entirely new work domain, often called the “envisioned world problem” [40, 41]. AI monitoring is a growing area that will likely result in new roles and responsibilities within health systems in the coming years. Because of this, there are no specific intended “users” of VAMOS to get design input from. To overcome this design challenge, we identified relevant analogues of AI monitoring tasks (e.g., CDS monitoring) and interviewed people in existing roles that use similar skills as those that will be needed for VAMOS (e.g., identifying anomalies, responding to health IT breakdowns). This was an approach used by Sushereba et al. [44] to design a first-of-a-kind rotorcraft for the Army. In line with the recommendation of Miller & Feigh [42], a large focus of the design process was spent defining the future work domain for VAMOS. This ensured the technology design supported the broader sociotechnical system of the envisioned work domain. Additional user research will be needed to refine the VAMOS system as new roles for AI monitoring emerge.

We adapted existing human factors design principles to incorporate the concept of workflow integration to our heuristic evaluation. Most human factors design principles traditionally used in heuristic evaluation focus on the user interface design, such as the grouping of information and use of terms. Yet, poor workflow integration of health technologies remains a persistent issue [52]. To try and address workflow integration in our design, our heuristic evaluation combined the design principles of Scapin and Bastien [46], Zhang et al. [47], and concepts of workflow integration based on the Checklist of Workflow Integration developed by Salwei et al. [48].

About half of the issues identified in the heuristic evaluation related to workflow integration concepts. For instance, we frequently discussed how VAMOS would support the next steps of the users, such as requesting a model consult or viewing the model data; these issues related to the principle “Flow of tasks: In sequence”. Other issues that arose in the heuristic evaluation included how VAMOS would support the work of the large AI monitoring team, for instance, indicating who on the team is assigned to a model in maintenance. This related to the workflow integration principle, “team level workflow”. These extended design principles can support the design of health technologies and may improve their integration in clinical workflow.

One limitation of this work is that we largely focused on monitoring of predictive AI models rather than generative AI. Future research should explore how to appropriately monitor generative AI applications [53]. As metrics develop for assessing generative AI model performance at scale, our flexible VAMOS design is well-positioned to incorporate these novel metrics. Another limitation is that we have not tested this system in the real world or across other diverse settings. We are currently building an early version of the VAMOS system as well as developing a common data model for algorithmovigilance systems. Next steps will include continual refinement and testing of the system with an initial set of four models currently in use at VUMC. We will then conduct robust usability testing to further refine the system followed by pilot testing. Throughout this process, we will continue to engage diverse stakeholders in the system design including input from clinical stakeholders, patients and their family caregivers.

Additionally, the value assumptions underlying such monitoring will also be more deeply investigated, so that the system can provide transparent rationales for key alerts. Future research should explore translating the system to other organizations, testing the system across diverse settings, and developing AI monitoring standards that enable inter-organizational sharing and reporting. This will enable algorithmovigilance and continuous learning across healthcare systems.

## CONCLUSION

The development of VAMOS represents a significant step forward in the ongoing effort to monitor and govern the use of AI within healthcare settings. By engaging a multidisciplinary team and involving diverse stakeholders through participatory design sessions and interviews, we identified design requirements and functionalities needed to support AI monitoring across its lifecycle. Our recommendations and insights can serve as a valuable guide for other institutions seeking to implement similar systems. VAMOS contributes to the broader goal of finding ways to ensure patient safety and maintaining trust in AI technologies in healthcare by enabling systematic and proactive monitoring, laying the groundwork for enhanced algorithmovigilance practices in the future.

## Data Availability

The de-identified data produced in the present study are available upon reasonable request to the authors.

## ACKNOWLEDGEMENTS

This research was funded by Kaiser Permanente/The Gordon and Betty Moore Foundation through the Augmented Intelligence in Medicine and Healthcare Initiative (AIM-HI). This work was also supported in part by the Agency for Healthcare Research and Quality (AHRQ) grant K01HS029042 (Salwei). The content is solely the responsibility of the authors and does not necessarily represent the official views of The Gordon and Betty Moore Foundation or AHRQ.

